# Educational inequalities in statin treatment for preventing cardiovascular disease: cross-sectional analysis of UK Biobank

**DOI:** 10.1101/2020.06.11.20128116

**Authors:** Alice R Carter, Dipender Gill, Richard Morris, George Davey Smith, Amy E Taylor, Neil M Davies, Laura D Howe

**Author notes:** **Corresponding author:** Ms Alice R Carter, Oakfield House, Oakfield Grove, Bristol, BS8 2BN, @alicerosecarter. NMD and LDH contributed equally.

## Abstract

**Background:** The most socioeconomically deprived individuals remain at the greatest risk of cardiovascular disease. Differences in risk adjusted use of statins between educational groups may contribute to these inequalities. We explore whether people with lower levels of educational attainment are less likely to take statins for a given level of cardiovascular risk.

**Methods and findings:** Using data from a large prospective cohort study, UK Biobank, we calculated a QRISK3 cardiovascular risk score for 472 097 eligible participants with complete data on self-reported educational attainment and statin use (55% female; mean age, 56). We used logistic regression to explore the association between i) QRISK3 score and self-report statin use and ii) educational attainment and self-report statin use. We then stratified the association of QRISK3 score, and statin use by strata of educational attainment to test for an interaction. In this sample, greater education was associated with lower statin use, whilst higher cardiovascular risk (assessed by QRISK3 score) was associated with higher statin use in both females and males. There was evidence of an interaction between QRISK3 and education, such that for the same QRISK3 score, people with more education were more likely to report taking statins. For example, in women with 7 years of schooling, equivalent to leaving school with no formal qualifications, a one unit increase in QRISK3 score was associated with a 7% higher odds of statin use (odds ratio (OR) 1.07, 95% CI 1.07, 1.07). In contrast, in women with 20 years of schooling, equivalent to obtaining a degree, a one unit increase in QRISK3 score was associated with an 14% higher odds of statin use (OR 1.14, 95% CI 1.14, 1.15). Comparable ORs in men were 1.04 (95% CI 1.04, 1.05) for men with 7 years of schooling and (95% CI 1.08, 1.08) for men with 20 years of schooling. Linkage between UK biobank and primary care data meant we were able to carry out a number of sensitivity analyses to test the robustness of our findings. However, a limitation of our study is that a number of assumptions were made when deriving QRISK3 scores which may overestimate the scores.

**Conclusions:** For the same level of cardiovascular risk, individuals with lower educational attainment are less likely to receive statins, likely contributing to health inequalities.

**Summary:** *What is already known on this topic?:*

- Despite reductions in the rates of cardiovascular disease in high income countries, individuals who are the most socioeconomically deprived remain at the highest risk.
- Although intermediate lifestyle and behavioural risk factors explain some of this, much of the effect remains unexplained. What does this study add?

- For the same increase in QRISK3 score, the likelihood of statin use increased more in individuals with high educational attainment compared with individuals with lower educational attainment.
- These results were similar when using UK Biobank to derive QRISK3 scores and when using QRISK scores recorded in primary care records, and when using self-reported statin prescription data or prescription data from linked primary care records.
- The mechanisms leading to these differences are unknown, but both health seeking behaviours and clinical factors may contribute.

## Introduction

Despite reductions in cardiovascular morbidity and mortality in high income countries, the most socioeconomically deprived groups remain at the highest risk of cardiovascular disease (CVD) (1, 2). There is evidence that education is a causal risk factor for CVD (3-5). We have previously demonstrated that part of this association acts through three modifiable risk factors; body mass index (BMI), systolic blood pressure and lifetime smoking behaviour (5). However, as much as 60% of the effect of education on CVD remains unexplained.

Previous studies have assessed the association of socioeconomic position (SEP) with primary (prescribed prior to a cardiovascular event) and secondary (prescribed as a result of a cardiovascular event) CVD preventative treatment rates; however, the direction of effect has been mixed (6-10). In an analysis of the Whitehall II cohort study (11), and in the British Regional Heart Study (12) there was no evidence of socioeconomic differences in statin prescribing. In other studies it has been reported that those with lower socioeconomic position are more likely to be prescribed statins (6, 8, 13, 14). Conversely, some studies have found that individuals of lower socioeconomic position are less likely to be prescribed statins (9, 10, 15, 16).

One key challenge in trying to unpick the role of education in statin prescribing (or other primary or secondary prevention mechanisms) is that lower education is associated with higher levels of cardiovascular risk factors. For example, lower education is associated with higher BMI, smoking, higher blood pressure, and lower levels of physical activity (5, 17, 18). Therefore, individuals with low education likely have a greater underlying risk of CVD and therefore potentially have a greater need for statins. However, it is possible that educational differences in health-seeking behaviour or interactions between patients and healthcare professionals may result in those with higher educational levels being prescribed preventative medication at a lower level of clinical ‘need’ (19, 20). Consequently, it is more informative to test whether there are educational differences in statin use dependent upon cardiovascular risk, rather than to look at the crude association of education and statin use.

Using the UK Biobank cohort, we investigated whether for a given level of cardiovascular risk, measured using the QRISK3 cardiovascular risk score, people with lower education were less likely to report taking statins as primary prevention than those with higher education (21-23). In secondary analyses we identify whether there are inequalities in the type of statin (Atorvastatin compared with Simvastatin) prescribed, given that Atorvastatin has greater efficacy than Simvastatin but is more costly (24-27).

## Methods

### UK Biobank

The UK Biobank study recruited 503 317 UK adults between 2006 and 2010. Participants attended baseline assessment centres involving questionnaires, interviews, anthropometric, physical and genetic measurements (15, 16). All UK Biobank participants are linked to hospital episode statistic (HES) data, with data available from 1997 in England, 1998 in Wales and 1981 in Scotland (28), with the most recent entry recorded in this analysis in May 2017. Additionally, a subset of participants (approximately 230,000 participants) are linked with primary care data and prescribing data (29). In this analysis, we use data from baseline assessment centres, HES data, and linked primary care data where available.

### QRISK risk score

We created a risk score for cardiovascular disease using the QRISK3 algorithm (23). The QRISK3 score is currently used in primary care systems in England and Wales to define the treatment threshold for statin prescriptions. Current guidelines recommend prescribing statins to individuals with a 10% or greater risk of having a cardiovascular event within 10 years (30, 31). QRISK3 scores were derived for all participants with complete data for educational attainment and reported statin use (N= 472 097) (Supplementary Figure 1). For individuals with missing data in any of the QRISK3 variables multiple imputation was used (see statistical analysis section). Scores were derived according to the publicly available QRISK3 algorithm https://qrisk.org/three/index.php.

**Figure 1:**
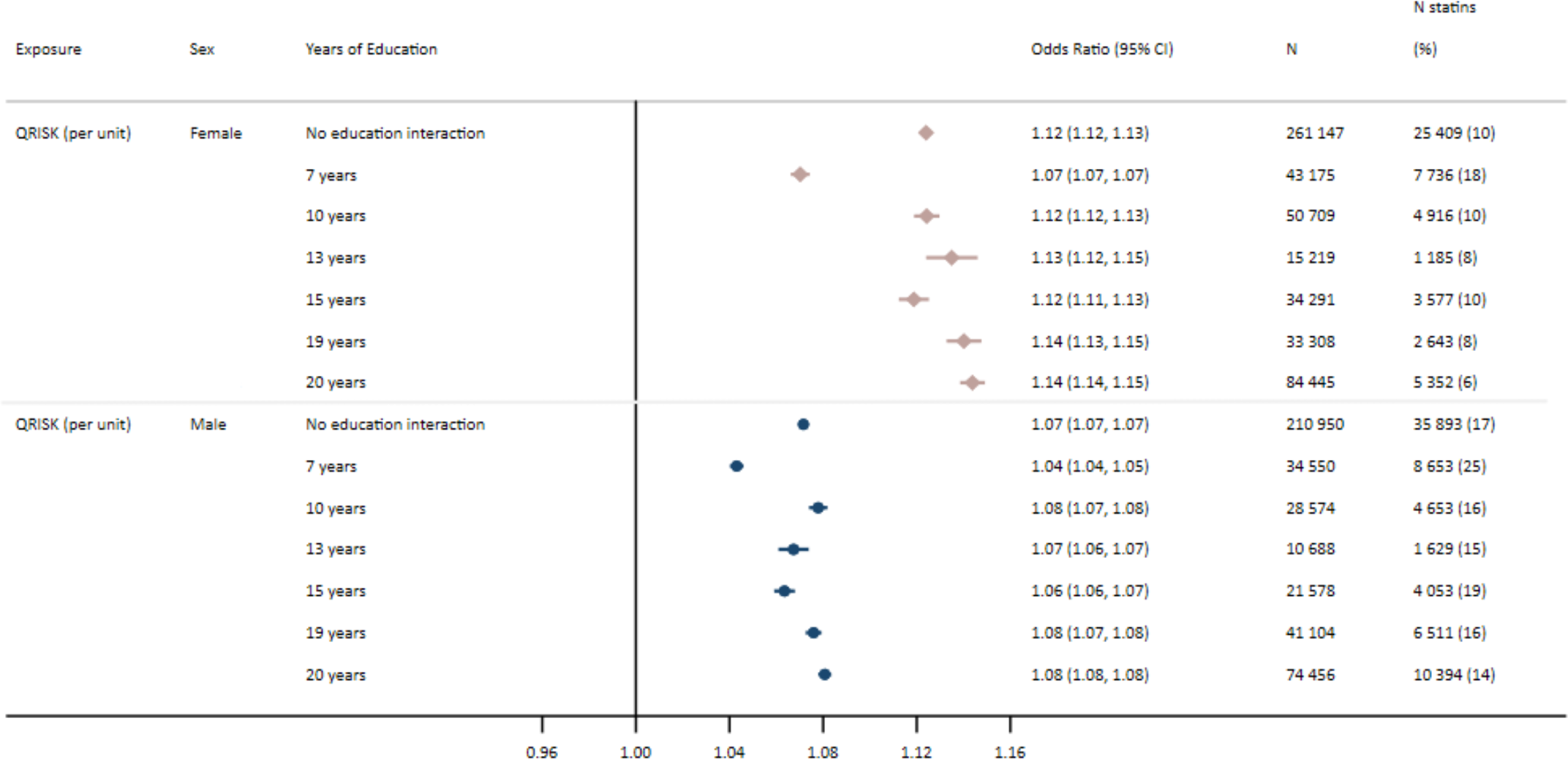
Odds of self-report statin use per unit increase in baseline QRISK3 score with no education interaction and stratified by years of education in females and males

Where measures were recorded in baseline assessment centres, such as BMI, Townsend deprivation score or systolic blood pressure, these values were used. With the exception of systolic blood pressure variability (standard deviation of repeated values) and coronary heart disease in a first-degree relative under 60 years of age, all QRISK3 variables were available in UK Biobank. In the absence of repeated measures of systolic blood pressure on UK biobank a measure of systolic blood pressure variability was derived from the standard deviation of the two recorded measurements of systolic blood pressure at the baseline assessment centre. A measure of family history of CVD was ascertained from reported heart disease in mothers, fathers and siblings of UK Biobank participants, however age of diagnosis and subtype of CVD, could not be determined.

Full details of all variable measurements are available in the supplementary materials. All variables used and assumptions made when deriving QRISK3 scores are available in Supplementary Table 1. UK Biobank treatment codes used to identify cases and ICD9, ICD10 codes and are presented in Supplementary Tables 2 and 3.

In a subset of individuals with linked primary care data, QRISK (read 2 code: 38DF.) (N=1 495) (21), or QRISK2 scores (read 2 code: 39DP.) (N = 10 633) (22) were recorded from 2007 onwards. Where more than one QRISK score was recorded for an individual, the first recorded value was used in analysis.

### Measuring educational attainment

UK Biobank participants reported their highest qualification achieved at baseline assessment centres, which was converted to the International Standard Classification for Education (ISCED) coding of years of education (Supplementary Table 4) (32).

### Measuring statin use

Participants were asked about regular medication they were taking, details of which were recorded by UK Biobank study nurses. From this, a primary variable for any reported statin use was generated. The type of statin used (Atorvastatin, Simvastatin, Fluvastatin, Pravastatin and Rosuvastatin) was recorded by study nurses and was used to derive a variable for type of statin.

In individuals with linked primary care data, statin prescriptions were recorded in prescription data. In these individuals, a measure of validated statin use was created, defined by a prescription in both the 3 months before and 3 months after baseline. For sensitivity analyses in individuals with a QRISK or QRISK2 score recorded in primary care data, statin use was defined as any statin prescription after a QRISK score was recorded.

### Exclusion criteria

Prevalent CVD cases at baseline, who would receive a statin prescription according to NICE guidelines (30, 31, 33, 34), were excluded from analyses. These cardiovascular diagnoses and events were ascertained through linkage to HES data, with cases defined according to ICD-9 and ICD-10 codes (Supplementary Table 3). Individuals were excluded if they had experienced at least one diagnosis of myocardial infarction, angina, stroke, transient ischaemic attack, peripheral arterial disease, type 1 diabetes, chronic kidney disease or familial hypercholesterolaemia (30, 34). The date for each diagnosis is provided in the HES data, which was linked with the date of assessment centre visit provided by UK Biobank.

Complete case analyses were carried out on 368 721 individuals, with complete data on age, sex, educational attainment, self-reported statin (medication) use and all variables required for the QRISK3 score (Supplementary Table 1 and Supplementary Figure 1).

### Code and data availability

The derived variables have been returned to UK Biobank for archiving. The code used to derive QRISK3 scores and carry out analyses is available at github.com/alicerosecarter/statin_inequalities.

### Patient and public involvement

Ethical approval for this study was sought from the UK Biobank (project 10953). No patients or participants were involved in setting the research question or the outcome measures, nor were they involved in developing plans for design or implementation of the study. No patients were asked to advise on the interpretation or writing up of results.

### Statistical analyses

To maximise power and potentially reduce bias, multivariable multiple imputation by chained equations (35) was used to impute variables included in the QRISK3 score with missing data, under the missing at random assumption. The sample for imputation was defined as all individuals with complete data on educational attainment and reported statin use. The proportion of missing data ranged from 0% to 15% (see Supplementary Table 5 for full details). In total, 77% of participants had no missing data, 13% of participants were missing data for one QRISK3 variable, 8% of participants were missing data for two QRISK3 variables and 2% of participants were missing data for three, four or five variables. A total of 25 imputed datasets were generated (36). Imputation was carried out separately within strata of years of education and sex to preserve interactions tested in the statistical analyses (37). The mean and standard deviation of continuous variables or proportion and standard error of categorical variables in the imputed data were compared with those from the complete data. All analyses were then carried out in each imputed dataset, with results combined according to Rubin’s rules.

It was determined *a priori* to carry out all analyses stratified by sex given the known differences in cardiovascular risk profiles for males and females (38, 39), as well as the QRISK3 score being derived separately by sex (23).

To confirm the validity of the derived QRISK3 score, a univariable logistic regression model was used to assess the association of the risk score with i) self-reported statin use and ii) incident CVD.

We estimated the associations of years of education with i) QRISK3 score (using linear regression) and ii) statin use (using logistic regression).

#### Testing for interaction between QRISK3 score and educational attainment on statin use

Logistic regression was used to estimate the association of QRISK3 score with self-reported statin use, stratified by years of education, providing an estimate of interaction on the multiplicative scale (sFigure 2, Route 1). These analyses were not adjusted for any other covariates, assuming all relevant variables are incorporated into the QRISK3 score. Evidence of an interaction between QRISK3 score and years of education was evaluated in a linear model where the interaction term QRISK3×educational attainment was included in the regression model.

**Figure 2:**
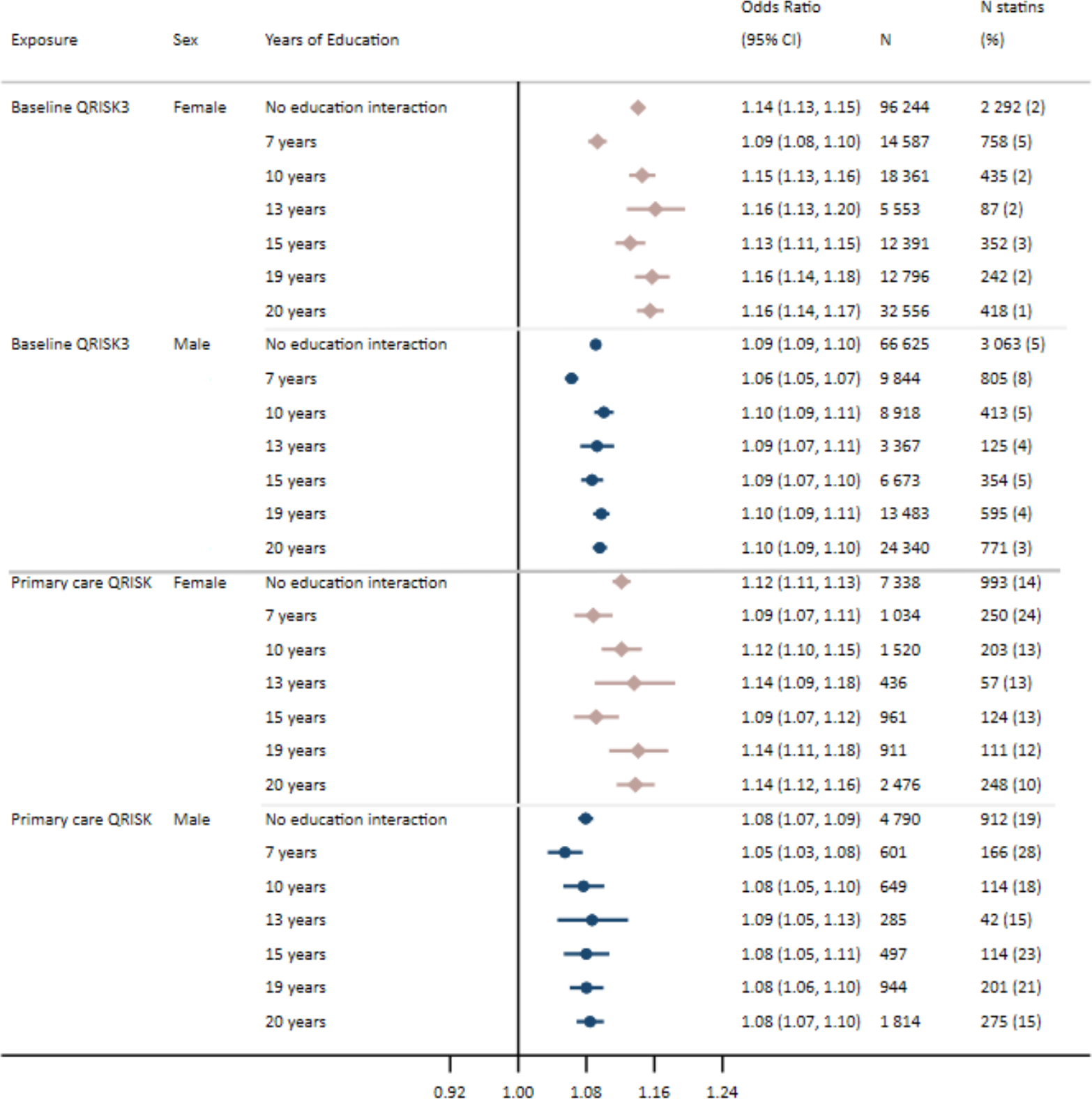
Odds of statin use recorded in primary care prescription data per unit increase in A) baseline QRISK3 score and B) QRISK or QRISK2 score recorded in primary care, in females and males

#### Secondary analyses

To test the hypothesis that there may be educational inequalities in the type of statin prescribed, in individuals who reported using statins to baseline study nurses, we assessed i) whether there was an association of QRISK3 score and years of education independently with self-reported Atorvastatin, which has been suggested to have a greater efficacy, compared with self-reported Simvastatin (baseline) (24-26) and ii) whether there was any evidence of an interaction between QRISK3 score and years of education on type of statin prescribed (sFigure 2, Route 2).

Analyses testing the association between QRISK3 and years of education on statin use and statin type independently, as well as for any interaction between QRISK3 score and educational attainment, on statin use and type of statin prescribed were replicated using complete case data (sFigure 2, Route 3 and 4).

To test whether the self-reported statin use data affected the results, we repeated our analysis with statin use defined as a prescription both 3 months before and after baseline from linked primary care prescription data (sFigure 2, Route 5), and also repeated our main analysis with self-reported statin use in the subset of participants with the linked prescription data (sFigure 2, Route 6).

In the subsample of primary care individuals with a QRISK or QRISK2 score recorded, analyses were replicated to test for evidence of an interaction between QRISK score and incident statin prescribing. This was defined as any prescription for a statin recorded in primary care data, excluding individuals who reported using statins to study nurses at the baseline assessment centre (sFigure 2, Route 7).

QRISK scores were included if they were recorded on or prior to the date of first statin prescription, but consideration was not given to the time between both events.

Two further estimates of QRISK3 were derived excluding i) variability of systolic blood pressure and ii) family history of cardiovascular disease from QRISK3 scores. The pairwise correlation between scores with and without these variables was tested.

## Results

### UK Biobank sample

In the main analyses (N = 472 097) 55% of participants were female with a mean age of 56. In females, the QRISK3 score implied a mean 10-year risk of a cardiovascular event of 6.9% (standard deviation (SD) = 5.5). In males, the QRISK3 score implied mean a 10-year risk of a cardiovascular even of 13.1% (SD = 8.4). Participants were more likely to have completed 20 years of education (female = 35%, male = 38%) than 7 years of education (female = 14%, male = 14%). 10% of females and 17% of males reported using statins.

The distribution of variables was similar between the multiply imputed dataset, complete case data, and in the subset of participants with linked primary care data (Supplementary Table 6).

### Association of QRISK3 score with statins and cardiovascular disease

For a one unit increase in QRISK3 score (i.e. a 1% increase in the 10-year risk of experiencing a cardiovascular event) in females, the odds ratio (OR) for reporting statin use to study nurses was 1.12 (95% confidence interval (CI): 1.12 to 1.13) and the OR for an incident cardiovascular event was 1.12 (95% CI: 1.12 to 1.12) (Figure 1 and Supplementary Table 7). Females with a QRISK3 score of 10 or greater were 1.34 (95% CI: 1.31 to 1.36) times more likely to report using statins than those with a QRISK score of less than 10. In males, the OR for statin use was 1.07 (95% CI: 1.07 to 1.07) and for an incident cardiovascular event the OR was 1.08 (95% CI: 1.08 to 1.08) per unit higher QRISK3 score (Figure 1 and Supplementary Table 7). Males with a QRISK3 score of 10 or greater were 1.49 (95% CI: 1.46 to 1.52) times more likely to report using statins than those with a QRISK score of less than 10.

#### Analyses stratified by years of education provide an estimate of interaction on the multiplicative scale

*P value for interaction in females = <0*.*001 and males = <0*.*001*

### Association of education with QRISK3 score and statin prescribing

Per year increase in educational attainment was associated with a −0.30 (95% CI: −0.30 to −0.29) reduction in mean QRISK3 score in females and a −0.35 (95% CI: −0.35 to −0.34) reduction in mean QRISK3 score in males (Supplementary Table 8 and Supplementary Figure 3).

The prevalence of statin use was highest in those in the lowest strata of educational attainment (equivalent to leaving school after 7 years, with no formal qualifications) (Supplementary Figure 4). Not accounting for cardiovascular risk, each additional year of education was associated with a lower odds of being prescribed statins (all types), (OR in females: 0.93; 95% CI: 0.93 to 0.93, OR in males: 0.96; 95% CI: 0.96 to 0.96) (Supplementary Table 7 and Supplementary Figure 5).

### Interaction between education and QRISK3 score in relation to statin prescribing

In both females and males, there was evidence of an interaction between QRISK3 score and years of education on statin use, such that for the same increase in QRISK3 score, the likelihood of statin use increased more for those of high educational attainment. In females, per unit increase in QRISK3, the OR for reporting statin use in those with the greatest years of education (20 years, equivalent to obtaining a degree) was 1.14 (95% CI: 1.14 to 1.15) compared with an OR of 1.07 (95% CI: 1.07 to 1.07) for those with the least years of education (7 years, equivalent to leaving school with no formal qualifications) (Figure 1). In males, the OR for statin use per unit increase in QRISK3 score in those with 20 years of education was 1.08 (95% CI: 1.08 to 1.08) compared with an OR of 1.04 (95% CI: 1.04 to 1.05) for those with 7 years of education (Figure 1). For both females and males, the P value for interaction was <0.001.

### Secondary analyses

Among individuals prescribed with either atorvastatin or simvastatin, those with higher QRISK3 scores were more likely to have been prescribed the more effective Atorvastatin. The OR for a one-unit higher QRISK3 and reporting Atorvastatin use was, OR: 1.02 (95%CI: 1.02 to 1.03) (Supplementary Table 9). This was similar in males, OR: 1.02 (95% CI: 1.01 to 1.02). Females, but not males, were less likely to have been prescribed Atorvastatin if they had more years of education; e.g. the OR for Atorvastatin prescription for 20 years of education versus 7 years of education was 0.92 in females (95% CI 0.83 to 1.01) and 1.02 in males (95% CI 0.94 to 1.11). There was little evidence of an interaction between QRISK3 score and educational attainment on statin type in females and males (P value for interaction in females = 0.4; P value for interaction in males = 0.9) (Supplementary Figure 6).

When analyses were replicated using eligible participants with linked primary care data using i) baseline measures of QRISK3 and self-report statin use, ii) baseline measures of QRISK3 with statin use validated by a prescription and iii) QRISK or QRISK2 score recorded in primary care data with a statin prescription, the evidence for interaction between QRISK3 and educational attainment on statin use remained in females (Figure 2 and Supplementary Figure 7). In males, the interaction between baseline QRISK3 scores and educational attainment on validated prescription remained. However, there was less evidence of an interaction between the primary care recorded QRISK scores and educational attainment on statin prescriptions (P=0.09), although the direction of effect was similar where males with 20 years of education were more likely to be prescribed statins (OR: 1.08; 95% CI: 1.07 to 1.10) than those with 7 years of education (OR: 1.05; 95% CI: 1.03 to 1.08) (Figure 2 and Supplementary Figure 7).

#### Analyses stratified by years of education provide an estimate of interaction on the multiplicative scale

*Baseline QRISK3: P value for interaction in females = <0*.*001 and males = <0*.*001*

*QRISK score recorded in primary care: P value for interaction in females = 0*.*034 and males = 0*.*091*

In the complete case sample, there was evidence of an interaction between QRISK3 and education in both males and females considering reported statin use as the outcome, where the P value for interaction was <0.001 for both females and males. However, there was little evidence of an interaction between QRISK3 and education on statin type (Supplementary Table 10).

Pairwise correlation between the baseline derived QRISK3 score and QRISK3 scores derived excluding i) systolic blood pressure variability estimated from the difference between two baseline measures and ii) self-report of any CVD in a mother, father or sibling, were high (all >0.97) (Supplementary Table 11).

## Discussion

Despite there being a higher prevalence of statin prescribing overall in those with lower levels of education, at a given level of QRISK3 score as a measure of clinical assessment of cardiovascular risk, less educated individuals were less likely to receive statin treatment compared to more highly educated individuals.

## Results in context

Lifestyle and behavioural factors, such as BMI, diet, smoking, risky drinking and exercise have previously been implicated as mediators of the association between education and CVD (17, 18, 40-46). Indeed, the higher overall prevalence of statins in lower educated individuals is likely due to the greater prevalence of these intermediate risk factors, compared with those of greater education (5, 17, 44). However, much of the association between education and CVD remains unexplained. The results presented in this analysis suggest that access to preventative medication for CVD may be contributing to persisting socioeconomic inequalities.

It has previously been reported that inequalities exist in favour of those with higher SEP when accessing preventative healthcare (47). In the UK, National Health Service (NHS) health checks are offered to all residents aged between 40 and 74 without pre-existing conditions every 5 years, with the aim of preventing a number of diseases including CVD (such as by calculating QRISK scores), kidney disease and dementia (48). In a recent systematic review by Bunten and colleagues, seven studies were identified that indicated uptake of these health checks is lower in more socioeconomically deprived groups (49). Additionally one study included in this systematic review identified a trend towards lower uptake in smokers; an important risk factor for CVD that is also socially patterned (49, 50). Similar findings were also reported by Wilson and colleagues (51). These reasons for non-uptake of health checks, in combination with the inequalities identified in this study, indicate that methods to improve engagement with NHS health checks and preventative screening methods may reduce inequalities in cardiovascular outcomes.

Indeed, differences in health seeking behaviours may be driving some of the inequalities in statin use identified in our study. However, when interaction analyses were repeated using QRISK or QRISK2 scores recorded in primary care data and primary care records of statin prescriptions, these inequalities remained. Therefore, attendance to primary care clinics cannot be the sole driver of these inequalities.

The literature is mixed in the direction to which inequalities in access to statins exist, where some studies suggest that individuals with lower socioeconomic position are less likely to be prescribed statins (9, 10, 15, 16) and other studies finding the opposite or no differences (6, 8, 11, 13, 14). Of these previous studies, there was limited consideration for underlying cardiovascular risk in the analyses. Some studies adjusted for cardiovascular comorbidities (8, 13, 15, 16) such as cholesterol level, diabetes status or prevalent cardiovascular events. However, we only identified one other study that comprehensively adjusted for cardiovascular risk (11). Forde and colleagues established risk status via 10-year absolute risk of coronary heart disease determined using the Framingham study (11, 52) and assessed SEP by British civil service grade of employment. In contrast to our results using educational attainment as a measure of SEP, they did not find evidence of inequalities in statin use. The differences in our results compared with Forde and colleagues could be the different measure of SEP used (income vs education) or due to cohort differences, where Forde and colleagues used an occupational cohort study and here, we have used a population-based cohort. Additionally, it has been demonstrated that the QRISK score has a greater predictive power compared with the Framingham score (53). Therefore, our analyses may better account for underlying differences in cardiovascular risk.

Currently, the QRISK3 scores capture the prevalence of key risk factors in individuals, such as BMI, blood pressure and smoking, but our results show that accounting for these factors alone is not enough to address cardiovascular inequalities. Cardiovascular risk scores may need to be adapted to pay greater attention to SEP; something that has been described previously in the literature (54-56) These risk scores should be in principle, easy to use and clear for clinicians, where it has previously been reported that the use of risk scores in general practice is a source of confusion (57).

Despite there being almost 30 000 first instances of statin prescriptions after 1^st^ January 2008 (where QRISK scores were first introduced in 2007), in the primary care data linked with UK Biobank, there were only around 14 000 individuals with a recorded QRISK or QRISK2 scores in the same data. This is higher than in previous research by Finnikin and colleagues, where they identified using primary care records, that only 27% of patients prescribed statins had a recorded QRISK2 score (58). However, the lack of recorded QRISK scores, suggests the decision to prescribe statin treatment may be independent of an objective measure of cardiovascular risk, and potentially prescribed based on more subjective measures by the clinician or the patient.

In individuals with linked primary care data, 14% of eligible participants reported using statins to study nurses, however only 3% of participants had a linked prescription in the three months before and after baseline. These individuals without a linked prescription are likely a combination of individuals who are purchasing statins over the counter, have received a prescription from a private clinician, or are no longer prescribed statins. The majority (91%) of those without a linked prescription reported taking Simvastatin (currently the only statin available as an over the counter medicine). Although we cannot definitively discern the reason these individuals did not have a prescription, it is possible that accessing statins through non-NHS GPs (i.e. through private practices) or over the counter is further contributing to inequalities in cardiovascular outcomes. There is, to date, little freely available data on the prevalence of purchasing statins over the counter, rather than via attending a primary care clinic. However, our data suggests it could indeed be highly prevalent in the population.

## Strengths and limitations

The major strength of our work is the large sample size and array of data available. Given the age range of our participants (45-76 years) reported statin use is highly prevalent (10% in females and 17% in males). Additionally, the linked primary care data for 44% of the eligible sample allowed us to i) validate self-reported statin use and ii) compare different mechanisms inequalities may be arising. Where inequalities are present in primary care recorded QRISK scores, inequalities are unlikely to be due to health seeking behaviour and more likely due to factors arising within clinic settings. Conversely, where data is used from UK Biobank baselines assessment, inequalities may be due to either differences in health seeking behaviour (i.e. attending NHS health checks) or factors that arise within the healthcare setting.

Lifestyle and behavioural characteristics, which are incorporated in to the QRISK3 score, are likely to be captured much more accurately and completely in UK Biobank compared with a primary care setting. However, there may be some settings where UK Biobank variables may have been measured differently than they would in primary care (59), such as non-fasting blood biomarker measurements. However, the magnitude to which these measurements differ is unlikely to introduce much bias to estimates of the QRISK3 score. Additionally, selection bias is present in UK biobank, where participants are generally of a higher socioeconomic position and healthier than the general population (60). Those who are of a lower socioeconomic position in the UK Biobank potentially differ to those of an equivalent socioeconomic position (or level of educational attainment) in the general population, where UK Biobank participants may be more health conscious and health aware. Therefore, it is possible that the inequalities in the wider population are greater than the inequalities reported here.

Despite our large sample size and wealth of data, a number of assumptions were made when generating the QRISK3 scores. For example, in the QRISK3 algorithm (23), the study authors specify medications should be considered if the individual has two or more prescriptions for each class of medication (e.g. corticosteroid or atypical antipsychotic). We were not able to determine the number of prescriptions at baseline and relied on a single self-report measure of medication use. Therefore, medication use may be overestimated in our sample, which would result in an overestimate of the QRISK3 score. Additionally, some measures, such as systolic blood pressure variability and coronary heart disease in a first degree relative under the age of 60, are not available in the UK Biobank data. Although we have included measures likely to capture some of these variables, this may introduce bias to the QRISK3 estimate in UK Biobank compared with a primary care setting.

The ISCED definitions of educational attainment (years in schooling) can differ with respect to other measures of socioeconomic position. For example, using ISCED definitions, individuals who left school with a vocational qualification are given a high number of years of schooling (19 years) but will typically go into manual labour jobs. This is likely to explain some of the non-linearities in effects stratified by educational attainment.

## Clinical implications

The results presented here highlight inequalities in statin use by educational attainment. Given the persisting inequalities in CVD, addressing the contribution of differences in statin prescription provides a clear policy target. The two complimentary data sources used in this analysis, UK Biobank baseline data and linked primary care data, indicate two potential mechanisms for these inequalities. Firstly, there are likely to be differences in health seeking behaviour such as in attending NHS health checks as previously evidenced in the literature. Secondly, the inequalities present in the primary care data suggest there are important interactions between the healthcare practitioner and patient that result in unequal prescribing of statins.

Healthcare professionals should consider potential biases in prescribing preventative treatments, or in carrying out risk assessments, such as calculating a QRISK score. Additionally, patient preference for treatment may be socially patterned (61). However, addressing these inequalities requires systemic change and different interventions may be required to address the different mechanisms of inequalities. For example, policy makers and healthcare professionals should consider how they can improve the uptake of NHS health checks, where these risk assessments are carried out, in those who are socioeconomically deprived.

## Conclusions

Our analyses demonstrate that at a given level of cardiovascular risk, people with lower levels of educational attainment are less likely to be prescribed statins than people with higher educational attainment, meaning differences in statin prescribing likely contribute to inequalities in cardiovascular disease. Policies should consider how these inequalities can be minimised.

## Contributions

ARC designed the study, cleaned and analysed the data, interpreted the results, wrote and revised the manuscript. DG advised on defining medications, interpreted the results and critically reviewed and revised the manuscript. RM advised on analyses, interpreted the results and critically reviewed and revised the manuscript. GDS, AET, NMD and LDH all designed the study, interpreted the results, critically reviewed and revised the manuscript and provided supervision for the project. NMD and LDH contributed equally and are joint senior authors on this manuscript. ARC and LDH serve as guarantors of the paper. The corresponding author attests that all listed authors meet authorship criteria and that no others meeting the criteria have been omitted.

## Transparency statement

ARC affirms that the manuscript is an honest, accurate and transparent account of the study being reported and no important aspects of the study have been omitted.

## Acknowledgements

This work was carried out using the computational facilities of the Advanced Computing Research Centre - http://www.bris.ac.uk/acrc/ and the Research Data Storage Facility of the University of Bristol - http://www.bris.ac.uk/acrc/storage/.

